# The Impact of Early Life Stress on the Genetic Influence on Brain and Cognitive Development in Children

**DOI:** 10.1101/2021.12.27.21268445

**Authors:** Hee-Hwan Wang, Seo-Yoon Moon, Hyun-Jin Kim, KaKyeong Kim, Woo-Young Ahn, Yoonjung Yoonie Joo, Jiook Cha

## Abstract

Early life stress (ELS), such as *abuse*, neglect, and maltreatment, is a well-known risk factor for mental illness. However, it is unclear how ELS affects the brain and cognitive development. Identifying specific relationships of ELS with the genetic and brain-related underpinnings of cognitive development may reveal biological mechanisms responsible for the negative impact of ELS and those that lead to individual differences in sensitivity (or resilience) to ELS. In this study, to investigate the interlinked processes of cognitive development, we analyzed the multimodal data of DNA genotypes, brain imaging (MRI), and neuropsychological assessment (NIH Toolbox) outcomes of 4,276 children (ages 9 to 10 years, European ancestry) from the Adolescent Brain Cognitive Development (ABCD) study. We estimated the genetic influence on cognitive capacity using genome-wide polygenic scores (GPSs). Our regression and mediation analyses revealed significant causal relationships for the gene-brain-cognition pathway: Brain structural development significantly mediated the genetic influence on cognitive development (*partial mediation effect = 0*.*016, P*_*FWE*_*<0*.*001*). Interestingly, within the triangular relationship, we found a significant moderation effect of *abuse* only on the gene-to-brain pathway (*Index of Moderated Mediation = −0*.*007; 95% CI= −0*.*012 ∼ −0*.*002; P*_*FWE*_*<0*.*05*). These findings indicate the negative modulatory effects of ELS on the genetic influence on brain structural development that lead to disadvantageous neurocognitive development in prepubertal children.

## Introduction

Early life stress (ELS), such as *abuse* and *neglect*, is a well-known major risk factor for maladaptive cognition, behaviors, and psychiatric disorders^1–4^ with long-term sequelae^5^. ELS modulates the expression of the genes responsible for stress physiology, emotion regulation, cognitive control, and learning and memory^6–9,10^. Animal literature shows a causal effect of ELS on cognition and behaviors^11–20^ and the epi-/genetic mechanisms within the brain^21–24^. In humans, similar mechanisms may be involved in psychopathology^25,26^, stress physiology^7^, or emotion regulation^27–29^.

Although extensive literature shows associations of ELS with poor cognitive outcomes and brain functional and structural representations^1,3,8^, exactly how ELS affects cognitive capacity in humans remains unclear. Moreover, the impact of ELS on the development of the brain and cognition in children remains unknown. Addressing these questions will provide much-needed insight into who, when, and where to intervene regarding the negative neurocognitive outcomes of ELS. Considering the well-known impact of ELS on cognition^3,10^, we hypothesize that ELS is likely to modulate genetic influences on brain development as well as cognitive development^3,5^.

Cognitive development involves multiple dynamic processes that are regulated by the complex interplay of genetic and environmental factors^30–32^. Despite its significant impact, however, less is known about the biological mechanisms of how early environmental factors such as ELS, interact with genetic influences and impact cognitive development in children. The recent development of the genome-wide polygenic score approach (GPS) allows individual genetic loadings for cognitive capacity (e.g., accounting for up to 13% of the variance in cognitive outcomes in adults^33^) to be estimated quantitatively. Using this rigorous computational approach, we aimed to test whether ELS modulates the gene-brain-cognition development process.

This study had two main questions. First, does a gene-brain-cognition pathway exist during developmental periods? If so, does ELS negatively affect the causal pathway? We investigated these questions using multimodal data, including genetics, brain imaging, psychological data, and cognitive testing in large samples of prepubertal children (ages 9 to 10 years) with a wide range of sociodemographic characteristics.

## Methods

### Study Participants

Participants were enrolled in the Adolescent Brain Cognitive Development (ABCD) study, an ongoing longitudinal multisite study of brain development and health of youth in the United States. We used the deidentified neuroimaging, demographic, genetic, and behavioral data from the ABCD 2.0 Data Release^34^, these data were collected across 21 sites from 2015 to 2018. The participants were 11,875 children from the U.S. who were aged 9-10 years^35^. After removing participants who had at least one missing value from genetic, neuroimaging, and ELS data, 4,276 participants were included in our analyses.

### Genotype data for Genome-wide polygenic scores (GPS)

Saliva samples were collected from study participants and were genotyped using the Affymetrix NIDA Smokescreen array (733,293 SNPs) at the Rutgers University Cell and DNA Repository. We removed any inferiorly genotyped SNPs with thresholds of (i) genotype call rate<95%, (ii) sample call rate<95%, and (iii) rare variants with minor allele frequency (MAF)<1%. We imputed the data using the Michigan Imputation Server^36^ based on the 1000 Genome phase 3 reference panel with Eagle ver2.4 phased output^37^. For the imputed 12,046,090 SNPs, we additionally removed data from any individuals with >5% missing genotypes; extreme heterozygosity (F coefficient > 3 standard deviation from the population mean); and SNPs with <0.4 imputation INFO score, >5% missingness rate, <1% MAF and Hardy-Weinberg equilibrium (p<10^−6^). Genetic ancestry was determined with the fastSTRUCTURE algorithm^38^, available from ABCD release 3.0. Our analysis was restricted to individuals with European ancestry to control for population stratification. We estimated both kinship coefficients (K.C.s) and principal components (P.C.s) to control familial relatedness and ancestry admixture using PC-Air^39^ and PC-Relate^40^. We selected unrelated samples that were inferred to be more distant than 4th-degree relatives (K.C.>0.022) and removed any genetic outliers that fell significantly outside (>6 S.D. limits) the center in P.C. space. In the rest of this paper, we used genotype data (11,301,999 variants) of 8,523 unrelated European samples after Q.C., and the first ten ancestrally informative P.C.s were used as covariates in all the reported analyses (**Supplementary Figure 1**).

To assess the subject-level genetic propensity for cognitive ability, we computed genome-wide polygenic scores (GPS) for cognitive performance (CP) and educational attainment (EA) using publicly available GWAS summary statistics based on the European population^30^. The GPSs were constructed using PRSice2^41^, adjusting for the first ten principal components (P.C.s) of the genotype data to control for population stratification.

### Brain Imaging-Anatomical Imaging

T1-weighted (T1w) 3D structural MRI images were acquired in the ABCD study. We processed the images following established protocols^42,43^: The gradient nonlinearity distortion correction method was performed on structural MRI scans to improve geometric accuracy and image intensity reproducibility^44^. Based on tissue segmentation and sparse spatial smoothing, intensity nonuniformity was corrected. Then, the data were then resampled with 1 mm isotropic voxels into rigid alignment with an atlas-derived brain.

Cortical surface reconstruction was applied using the following procedures: structural MRI scans were processed using FreeSurfer v6.0 (https://surfer.nmr.mgh.harvard.edu) for cortical surface reconstruction^45^, which includes skull-stripping^46^, white matter segmentation and initial mesh creation^45^, correction of topological defects, surface optimization^47,48^, and nonlinear registration to a spherical surface-based atlas^49^.

### Brain Imaging-Diffusion Spectrum Imaging

We used the diffusion spectrum images from the ABCD study that were preprocessed using the following protocol^50^ by the ABCD Data Analysis and Informatics Center (DAIC). Eddy current distortion correction was used with a nonlinear estimation using diffusion gradient orientations and amplitudes to predict the pattern of distortion^51^. Head motion was corrected by registering images synthesized from tensor fit^52^. Diffusion gradients were adjusted for head rotation^52,53^. To identify and replace dark slices due to abrupt head motion, we used robust diffusion tensor estimation^54^. B0 distortion was corrected with the reversing gradient method^55^. Gradient nonlinearity distortion correction was applied^44^. The data were resampled to a standard orientation with an isotropic resolution of 1.5 mm.

To estimate accurate brain imaging phenotypes, we used individual connectome data. We applied MRtrix3^56^ for whole-brain white matter tract estimation and individualized connectome generation. For connectivity metrics, we used streamline counts associated with fiber connection strength^57,58^ associated with fiber integrity. We decreased noise^59^, and performed bias correction with the Advanced Normalization Tools (ANTs) pipeline’s N4 algorithm^60^. To obtain a connectivity index with a white matter pathway^61^, we performed probabilistic tractography by second-order integration over fiber orientation distributions^62^, with random seeding across the brain and target streamline counts of 20 million. These initial tractograms were filtered from preliminary tractograms using spherical-deconvolution informed filtering (2:1 ratio). With a final streamline count of 10 million, we generated an 84 × 84 whole-brain connectome matrix for each participant using the T1-based parcellation and segmentation in FreeSurfer. This pipeline ensured that individual participants’ connectomes were restricted to their own neuroanatomy. We carried out the computation on the supercomputers at Argonne Leadership Computing Facility Theta and Texas Advanced Computing Center Stampede2.

### Dimensionality reduction of brain data

To extract brain morphometric representations correlated with genetic influence on intelligence, we first performed generalized linear model analysis using cognitive capacity GPSs as independent variables and neuroimaging data as dependent variables adjusted to covariates (e.g., age, sex, maternal education, income, BMI, study site, and marital status). Among three different types of neuroimaging data (structural MRI, DTI-count, and DTI-FA), only brain morphometric features (i.e., structural MRI) showed significant associations with cognitive capacity GPSs (*P*_*FWE*_ *< 0*.*05*; **Figure 3, Supplementary Figure 2, Supplementary Table 1**)^63^. We then performed principal component analysis to extract representations from brain morphological features associated with CP GPS or EA GPS (R version 3.4.1). We used the first principal components as representations of the brain.

### NIH Toolbox Cognition Battery data

NIH Toolbox Cognitive Function Tests^64^ were used to assess various levels of general cognitive ability of the participants. For each child, summary scores from the NIH Toolbox Cognition Battery were provided, including the crystallized intelligence composite score (mainly measuring prior learning and past experiences about language), the fluid intelligence composite score (mainly measuring abstract reasoning and learning ability in novel situations), and total intelligence composite score, which is the combination of both crystallized and fluid intelligence composite scores^65,66^. The crystallized intelligence composite score is a composite of the *Picture Vocabulary Test* and the *Oral Reading Recognition Test* outcomes, and the fluid intelligence composite score is a composite of the *Dimensional Change Card Sort Test*, the *Flanker Inhibitory Contro*l and *Attention Test*, the *Picture Sequence Memory Test*, the *List Sorting Working Memory Test*, and the *Pattern Comparison Processing Speed Test* outcomes. Total intelligence composite score is a representation of general intelligence and is an aggregation of all the tests^67^.

### Early life stress

We derived ELS measures based on child exposure domains in the ABCD study^68^ (**Supplementary Table 2**). ELS measures were divided into three main categories: *household challenges, neglect*, and *abuse*. Subcategories of ELS data were as follows: *parental separation or divorce, criminal household member, household substance abuse, mental illness in household, mother treated violently* in *household challenges, emotional neglect, physical neglect* in *neglect*; and *physical abuse* and *sexual abuse* in *abuse*. We extracted the items of each subscale from the following various measurement tools (participant-or parent-reported): ABCD Family Environment Scale-Family Conflict Subscale Modified from PhenX, ABCD Diagnostic Interview for DSM-5 Traumatic Events, ABCD Family History Assessment, ABCD Parent Demographics Survey, ABCD Children’s Report of Parental Behavioral Inventory, and ABCD Parental Monitoring Survey. We averaged the measurements for each subcategory and transformed them into z scores. The higher the score, the more stressful experiences children had.

### Statistical Analysis

We performed a generalized linear model for three types of relationships — GPS-brain, GPS-intelligence composite scores, and brain-intelligence composite scores. We included the following covariates into those models: age, sex, maternal education, parental income, BMI, study site, and marital status. Family ID was not included because our analysis was performed in the dataset of unrelated individuals after excluding any relatives fourth-degree or closer during the genetic QC process. We used familywise error (i.e., Bonferroni correction) in each type of relationship for multiple comparison corrections.

For path modeling, we first tested an initial mediation model (**Figure 1-a**) to check whether the key relationship of the GPS-brain-cognition pathway was significant. Before examining the role of ELS in the GPS-brain-cognition pathway, we tested whether ELS directly affects cognition and whether ELS moderates the gene-cognition pathway (**Figure 1-b**). After we determined the potential moderation effect of ELS on the GPS-brain-cognition pathway, we evaluated the first-stage moderated mediation models and the second-stage moderated mediation models, following the framework of moderated mediation analysis^69–71^. These models assessed how ELS and its interaction with genetics or the brain, would affect the triangular pathway. We repeated the analyses with different subtypes of ELS and compared their effects.

**Figure 1.**
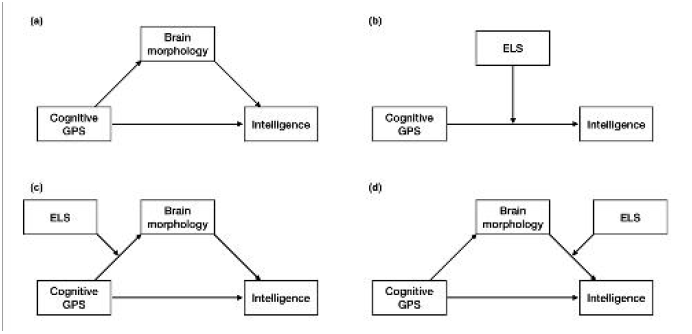
Schematic overview of the tested path models. (a) The baseline mediation model has the brain factor as a mediator, (b) The moderation model has ELS as a moderator, (c) The first-stage moderated mediation model for testing the moderating role of ELS on the relationship between the brain and genomic factors, (c) The second-stage moderated mediation model for testing the moderating role of ELS on the relationship between the brain and Intelligence composite scores.

Mediation and moderated mediation analyses were performed in R environment v3.4.1 using the *lavaan* v0.6-7 package with 1000 bootstrapping replicates. Mediation models included the same covariates that were used in the regression model. *Parental separation or divorce* had to be excluded in a moderated mediation model because of model convergence issues.

## Results

### Participants

After quality control, our study cohort comprised 4,276 unrelated children of European ancestry with a mean age of 9-10 years from the ABCD study^35^. **The** demographic characteristics of the patients, stratified by records of ELS experience, are summarized in **Table 1** (**Supplementary Table 3**) **and Figure 2**. All covariates, except for BMI, showed significant differences in experience with one or more type of ELS (*abuse: P < 0*.*001, neglect: P < 0*.*05, and household challenges: P < 0*.*05*) (**Supplementary Table 4**).

**Table 1.**
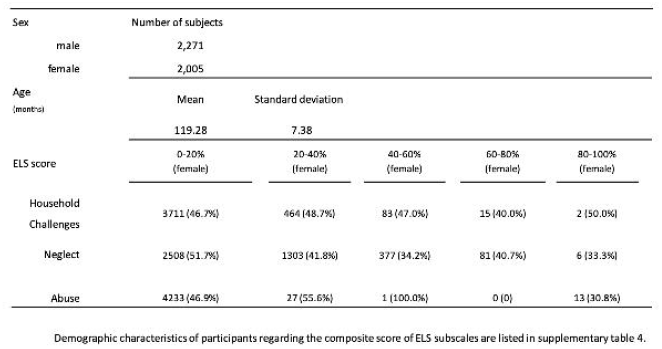
Demographic Characteristics of participants With European ancestry (N =4,276). The total participants are divided into five intervals regarding the Composite score of ELS.

**Figure 2.**
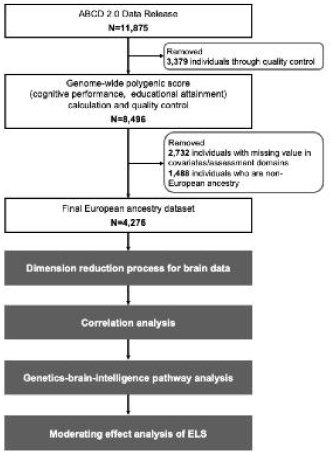
Flow chart of data selection and research design. From the ABCD 2.0 Data release, genetic, brain imaging, and cognitive assessment data for 11,875 participants were collected. For the 8,496 individuals after initial quality control and GPS calculalion, we additionally removed samples of non-European ancestry or with any missing values, for a total of 4,276 participants included in the analysis.

### Correlation among GPS, Brain, and Intelligence

#### Cognitive Capacity GPS-Brain

Out of 992 brain morphometric features, 169 and 44 brain features correlated significantly with CP GPS (*P*_*FWE*_*< 0*.*05*) and EA GPS, respectively (*P*_*FWE*_*< 0*.*05*), when adjusted for age, sex, maternal education, income, BMI, study site, and marital status (**Figure 3**). Of note, global brain features - including total gray matter volume, total cortex volume, subcortical gray matter volume, and total cerebral white matter volume - were shown to have significant associations with both GPSs (**Supplementary Table 1**). White matter fiber counts and fractional anisotropy of the structural connectomes showed no significant correlations with either GPS (**Supplementary Figure 2**). To obtain cognitive capacity GPS-related brain representations, we linearly reduced the dimensionality of brain morphometric features correlated with each GPS by performing principal component analysis. We verified that only the first components substantially explained the variance of each GPS-related brain morphometric feature (Brain _CP GPS_: 44.5%; Brain _EA GPS_: 54.9%) (**Supplementary Figurea 3, 4, 5**) and were significantly correlated with cognitive capacity GPS (Brain _CP GPS_: *β = 0.10, P*_*FWE*_*< 0.001;* Brain _EA GPS_: *β = 0.15, P*_*FWE*_*< 0.001*) (**Supplementary Table 5**).

**Figure 3.**
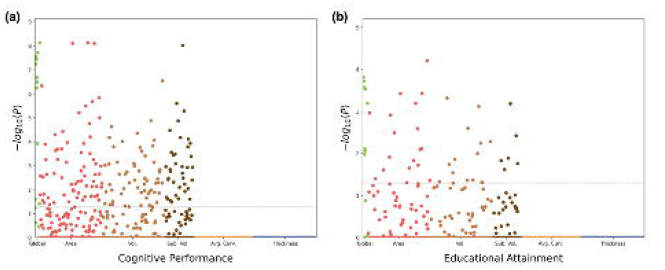
Manhattan plots of GPS-brain linear regression. The results of generalized linear models between brain morphometric features and (a) CP GPS and (b) between brain morphometric features and EA GPS are presented usi ng the Bonferroni-cor rected p values. The dotted line indicates the significance threshold(*P* _*FWE*_ =0.05). TheX-axis is the category of brain features: global features (Global), brain cortical area (Area), cortical volume (Volume), subcortical volume (Sub. Vol.), average cortical curvature(Avg. Curv), and cortical thickness (Thickness).

#### Brain-Intelligence Composite Score

Both *Brain* _*CP GPS*_ and *Brain* _*EA GPS*_ were significantly correlated with all three types of intelligence composite scores (*P*_*FWE*_*< 0.001*). Among the different types of intelligence composite scores, both brain representations showed the highest effect size on crystallized intelligence composite scores (*Brain* _*CP GPS*_: *β = 0.18; Brain* _*EA GPS*_: *β = 0.18*), with the lowest effect size on fluid intelligence composite scores (*Brain* _*CP GPS*_: *β = 0.05; Brain* _*EA GPS*_: β*= 0.05*). Total intelligence composite score was moderately correlated with brain representations (*Brain* _*CP GPS*_: *β = 0.12; Brain* _*EA GPS*_: *β = 0.12*) (**Supplementary Table 6**).

#### GPS-Intelligence Composite Score

Both CP and EA GPS were significantly correlated with the outcomes of all three of the different types of intelligence measures (*P*_*FWE*_*< 0.05)*. Test statistics showed that composite scores of crystallized intelligence appeared to have the strongest associations with both GPSs (*CP GPS:* β*= 0.17; EA GPS: β = 0.15*), followed by total intelligence composite scores (*CP GPS: β = 0.14; EA GPS: β = 0.10*) and the fluid intelligence composite score (*CP GPS: β = 0.09; EA GPS: β = 0.05*) (**Supplementary Table 6**).

### Mediation Analysis

Our analysis of the GPS-brain-cognition pathway (**Figure 1-a**) showed that the brain morphometric representations significantly mediated the causal effect of both GPSs on all types of intelligence (*P*_*FWE*_*< 0.001)*. Both GPSs showed the strongest direct effect on crystallized intelligence composite scores (β ***direct effect of CP GPS*** *= 0.15;* β ***direct effect of EA GPS*** *= 0.13)* compared to that on fluid intelligence composite scores (β ***direct effect of CP GPS*** *= 0.09;* β ***direct effect of EA GPS*** *= 0.04)* and total intelligence composite scores (β ***direct effect of CP GPS*** *= 0.13;* β ***direct effect of EA GPS*** *= 0.09)*. Both GPSs also showed the strongest indirect effect size on crystallized intelligence composite scores, which were determined by brain morphometric representation (β ***indirect effect of CP GPS*** *= 0.016;* β ***indirect effect of EA GPS*** *= 0.016)*, compared to fluid intelligence composite scores (β ***indirect effect of CP GPS*** *= 0.004;* β ***indirect effect of EA GPS*** *= 0.005)* and total intelligence composite scores (β ***indirect effect of CP GPS*** *= 0.010;* β ***indirect effect of EA GPS*** *= 0.010)* (**Table 2, Supplementary Table 7, Figure 4**).

**Table 2.**
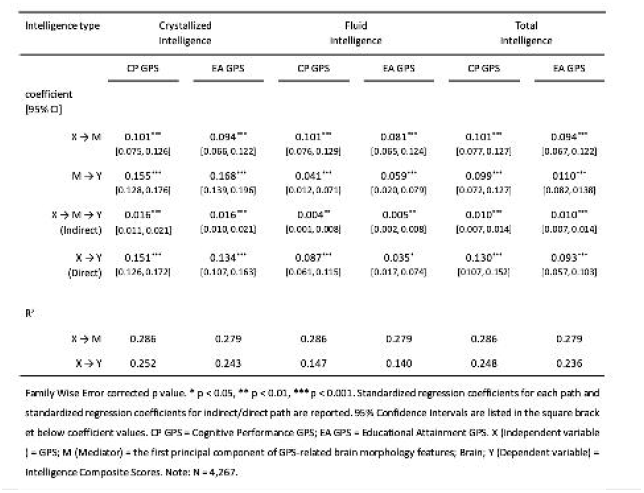
Results of mediation analysis.

**Figure 4.**
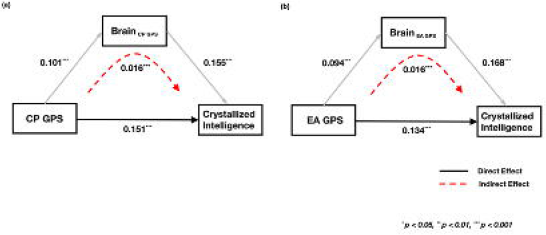
Path diagrams for crystallized intelligence mediation models. Standardizes regression coefficients for each path. Red dotted lines indicate the total indirect effect.

**Figure 5.**
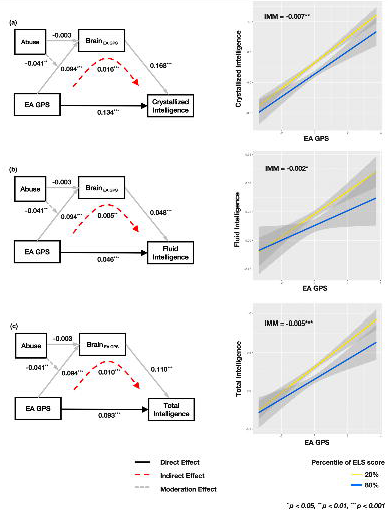
Effect of abuse on the EA GPS-brain-cognition pathway. The left panel shows standardized regression coefficients for each path. Red dotted lines indicate the total indirect effect. Gray dotted lines indicate the moderation effect of ELS on the GPS-brain relationship. The right panel shows differences in the indirect impact caused by composite ELS scores (transparent gray areas indicate confidence intervals).

#### Path Analysis: Moderated Mediation effect of ELS

Prior to moderated mediation analysis, we first assessed the correlation between ELS and intelligence composite scores and the evaluated the moderation effects of ELS on the relationship between cognitive capacity GPSs and intelligence composite scores (**Figure 1-b**). Among the different types of ELS, only *neglect* showed significant correlations with all types of intelligence composite scores (**Supplementary Table 8**). In moderation analysis, no significant moderation effects of the ELS variables were detected (*PFWE > 0.05*) (**Supplementary Table 9**).

The following first-stage moderated mediation analysis (**Figure 1-c**) showed the significant negative moderation effect of *abuse* on the EA GPS-brain-intelligence composite score relationship. Notably, *abuse* itself did not affect EA GPS-related brain morphometric representation; however, the interaction between EA GPS and *abuse* showed a significant negative association with brain morphometric representation (*β = −0.04*, ***95% CI*** *= −0.07 ∼ − 0.014, PFWE < 0.05)*. Among the different kinds of intelligence composite scores, crystallized intelligence composite scores were the most severely affected by the interplay of ELS and cognitive capacity GPSs through the brain (***Index of Moderated Mediation Crystallized Intelligence Score*** *= −0.007*, ***95% CI*** *= −0.012 ∼ −0.002; PFWE < 0.05*) compared to the effect on fluid intelligence composite scores (***Index of Moderated Mediation Fluid Intelligence Score*** *= − 0.002*, ***95% CI*** *= −0.004∼0.0; PFWE < 0.01*) and total intelligence composite scores (***Index of Moderated Mediation Total Intelligence Score*** *= −0.005*, ***95% CI*** *= −0.009 ∼ −0.002; PFWE < 0.01*) (**Table 3, Supplementary Table 10**). In the second-stage moderated mediation models (**Figure 1-d**), we found no significant interactions between the brain and ELS on intelligence composite scores (**Supplementary Table 11**). Taken together, the negative moderation effects of *abuse* on the causal pathway between genetic and phenotypic intelligence were only observed when the brain served as a mediator. The moderated mediation effect was only significant for *abuse*, but not for other ELS variables, even in subscales (**Supplementary Tables 12, 13**).

**Table 3.**
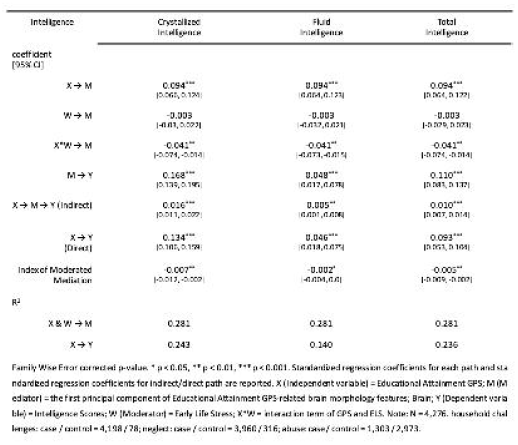
Moderated Mediation effect of *Abuse* on Educational Attainment GPS-Brain-Cognition.

## Discussion

By leveraging the multimodal genetic, neuroimaging, and cognitive assessment data of 4,267 children of European ancestry, we examined the complex interplay among brain structure and genomic influence on cognitive development and how ELS affects the gene-brain-cognition triad. Our analysis reveals likely causal neurodevelopment pathways under the genetic control of intelligence and ELS modulation. We found that brain structural development partially mediates the genomic contributions to cognitive development in young children. Of note, ELS (*abuse*) specifically modulates the influence of cognitive capacity GPSs on the brain, this modulatory effect then affects cognition. These results show the potential causal relationships between ELS (*abuse*) and the genetic pathway for neurocognitive development, accounting for biological how it may lead to cognitive deficits in children.

Extending the existing literature reporting the influence of genetics on the brain and cognition^3,72–77^, our study shows a more specific multifactorial model in which brain structural development mediates the genetic impact on cognitive development. Of note, this study shows that the association of the cognitive capacity GPS was only significant with gray matter phenotypes but not with white matter phenotypes. This result may be related to the developmental characteristics observed during preadolescence (9 to 10 years old), i.e., marked changes in gray matter, followed by relatively greater white matter changes or myelination^78^. Additionally, the brain mediation effect may be widespread involving several brain regions, considering that the brain mediator was the first principal component derived from hundreds of brain morphological features. Future research should examine how genetic influence is linked to longitudinal changes in brain structure over one’s lifetime.

The key contribution of this study to the literature is the discovery that ELS modulates the magnitude of the impact of genomic influence on neurocognitive development. This finding may account for a potential mechanism of the long-term effect of ELS on cognitive deficits. Note that our brain imaging results shows that ELS modulated the genomic influence on the cortical morphology of the various brain regions responsible for cognition and intelligence. In addition to the literature documenting the impact of ELS on affective processing, stress physiology and the relevant brain system^3,72–77^, this finding shows that ELS impacts cognitive brain circuitry.

Our moderated mediation results support genetic differential susceptibility to ELS. The genetic differential susceptibility model proposes that genetic variations of individuals regulate sensitivity or susceptibility to environmental influences^79–81^. In our analysis, as a result of the ELS moderation effects on the gene-to-brain pathway, children with the same level of ELS showed different cognitive outcomes depending on the magnitude of the EA GPS. That is, a child with a higher EA GPS showed a greater negative impact of ELS on the cognitive outcome, whereas one with a lower EA GPS showed a smaller negative impact.

Perhaps, in the case of individual with a high GPS, ELS may negatively regulate the expression of the genes responsible for the brain and cognitive development (sequentially as supported in our model). Consequently, otherwise, positive effects of high GPSs on cognitive outcomes may be eliminated under the effect of ELS, resulting in lower cognitive outcomes compared with the individuals with high GPSs and no ELS. A crucial outstanding question is whether this modulatory effect of ELS on the cognitive capacity GPS is linked to epigenetics. Animal research shows that ELS induces epigenetic modification in DNA methylation sites that are associated with a wide array of neural events, such as neurotransmitter biosynthesis, neurological system processes, glial cell proliferation, neurogenesis in the hippocampus, neural migration in the cerebral cortex, neuroplasticity, and neurodevelopmental delay^82–84^; many of these events are critically related to cognitive development. This line of research may allow precise risk stratification and the development of more individualized intervention strategies.

Another novel aspect of this study was the application of the GPS approach, rather than focusing on selected common variants of DNA, which permitted integrative assessment of the polygenic contribution to cognitive development and revealed its impact on the wide array of brain circuits. A recent study reported a similar finding of the association between cognitive capacity GPS and brain volumes in adult samples in the UK Biobank^73^. Our study showed that the gene-brain-cognition relationship manifests as early as preadolescence.

Our observations suggested that *physical or sexual abuse* significantly modulates the “gene-to-brain” path, which then leads to “cognition” but not the “gene-to-cognition” path. This finding is in line with previous ELS studies reporting that *physical neglect* and *abuse* negatively impact genetic regulation of brain systems, such as hippocampal glucocorticoid receptors, neurogenesis, and regulation of brain development^6,7,9,82^.

The direct effect of cognitive capacity GPSs on cognition was tenfold larger in magnitude than the indirect effects on the brain. Note that the gray matter representations derived from ROI-level morphometry may only partially account for the gene-to-brain effects. Indeed, a morphometric analysis may not fully detect subtle changes in the brain tissues under the control of epigenetic (or any other environmental) mechanisms, such as vascularization, neurogenesis, and synaptogenesis. Future research may elucidate sensitive brain representations related to genetic influence and environmental modulation.

Another possible explanation of the small indirect effect on the brain may be related to the limitations of linear models in testing complex nonlinear relationships. We used PCA to extract the brain representation (i.e., principal components) from thousands of brain morphometric variables. We used this method for the ease of statistical modeling (mediation analysis); however, since this method uses a linear and orthogonal transformation of data, we admit that this representation might be too simplistic to capture nonlinear relationships among the brain variables.

Our study showed the utility of our multitrait (cognitive performance and educational attainment) genomic approach for revealing the different patterns of genetic influences on the brain and cognition. Years of education (educational attainment, EA) is a widely used proxy for an intelligence phenotype. This is because of its high genetic correlation with intelligence and the ease of assessment compared to an evaluation of cognitive ability that would require behavioral tests (hence, it is unfavorable in large GWASs). For example, the literature shows that EA GPS correlates with cognitive functioning^85^ and the development of behaviors^86^.

Although both cognitive capacity GPSs showed significant effects on the gene-brain-cognition pathway, only EA GPS showed a significant modulatory effect of ELS on the pathway; CP GPS did not have a notable effect on the pathway. This observed ELS effect on EA GPS may be related to genetic loading not only for cognitive capacity but also for noncognitive skills and traits required for successful education attainment^87^. On the other hand, CP GPS correlated with a greater number of brain morphometric features than EA GPS did. This may reflect that CP GPS represents the genetic influence primarily on biological processes (i.e., brain development) directly linked to cognitive performance, but not so much on noncognitive processes. These points may be used to guide to future studies examining the gene-environment interaction in cognitive development.

Our results indicated greater magnitudes of the genetic influence on crystallized intelligence than on fluid intelligence during development. Crystallized intelligence is the ability to apply prior knowledge to problem solving; fluid intelligence is the ability to reason in novel situations without prior knowledge^65,88,89^. Since the genetic influence (both direct and indirect) was up to three times larger on crystallized intelligence compared to that on fluid intelligence, the impact of ELS via the gene-brain pathway was proportionally greater on crystallized intelligence than on fluid intelligence. This result adds granularity regarding the specificity of ELS impacts.

A few limitations of the study should be noted. First, it was not within the scope of this study to examine brain function, which is influenced by ELS through the triangular relationship. It is well known that ELS leads to changes in brain function^3,8^. Previous animal studies support that the function of adaptive gene-brain-cognition feedback systems may be affected by ELS^8^. Thus, the comprehensive impact of ELS on this triangular relationship needs to be further investigated in terms of brain function to reveal all the causal mechanisms. Second, we created composite scores of ELS with several proxy measures instead of using the established measure for childhood trauma, i.e., the Adverse Childhood Experiences (ACE) questionnaire^90^. This was mainly due to the unavailability of the ACE questionnaire data in the ABCD study. However, it is of note that we included questionnaires similar to those of the ACE questionnaire and included several ELS-related measurements^68^, thus aggregating various adverse environmental factors.

## Data Availability

All data used are available online at NIMH Data Archive

https://nda.nih.gov

## Acknowledgments

This work was supported by the New Faculty Startup Fund from Seoul National University and a Research grant from Basic Science Research Program through the National Research Foundation of Korea (NRF) funded by the Ministry of Education, South Korea (2021R1I1A1A01054995).

## Author contributions

- Study concept and design: Y.J., J.C.
- Acquisition, analysis, or interpretation of data: Y.J., H.W., S.M., K.K., H.K.
- Drafting of the manuscript: Y.J., H.W., S.M., J.C.
- Critical revision of the manuscript for important intellectual content: Y.J., J.K., H.W. S.M., K.K., W.A.
- Statistical analysis: H.W., S.M.
- Obtained funding: J.C.
- Study supervision: Y.J. J.C.

## Data and Code Availability

Codes and data are freely available for reproducibility (https://github.com/Transconnectome/ELS_on_genes-brain-cognition)

