## Supplementary Figures for "The Impact of Early Life Stress on the Genetic Influence on Brain and Cognitive Development in Children"

**Supplementary Figure**

**Supplementary Figure 1. Biplot from Principal Component Analysis on the combined genotype data of 1000Genome reference panel^1^ and the ABCD study participants (colored blue).** Super-populations are defined broadly as: AFR African, AMR admixed American, EAS East Asian, EUR European, SAS South Asian. To facilitate the understanding of genetic ancestry of the study participants, PCA was performed with 1000Genome phase3 reference panel and plotted with the first two principal components. Since ABCD participants reside on a continuum of genetic ancestry as American population (AMR), rather than distinct population groups, and none of them belongs to either African (AFR) or East Asian (EAS) populations, we did not exclude any genetic outliers based on the plot.
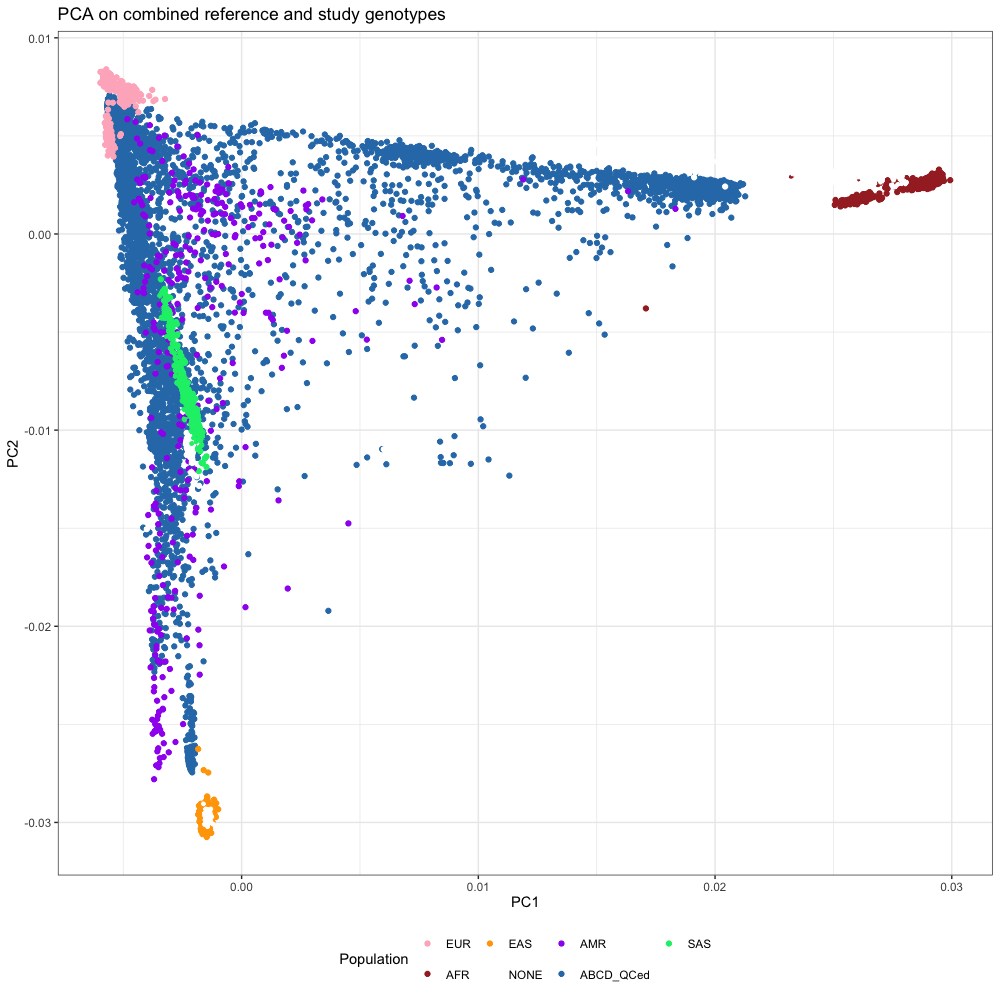

**Supplementary Figure 2.** **The result of generalized linear models between brain connectome and CP GPS (a, b), EA GPS (c, d).** The dotted line indicates the significance threshold (*P_bonferroni_* = 0.05).

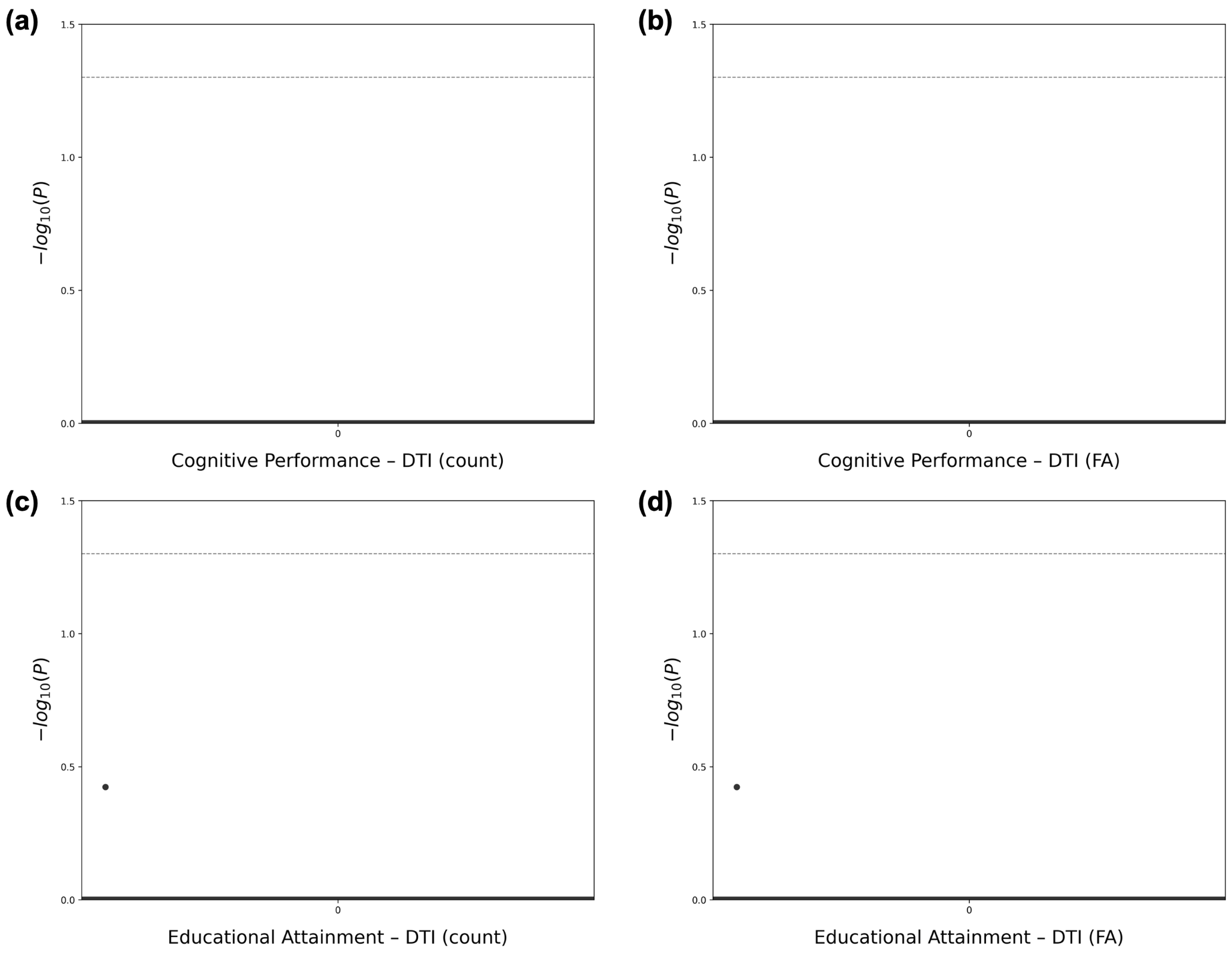

**Supplementary Figure 3. Proportion of explained variances by principal components from cognitive GPSs-related brain morphometric features.**

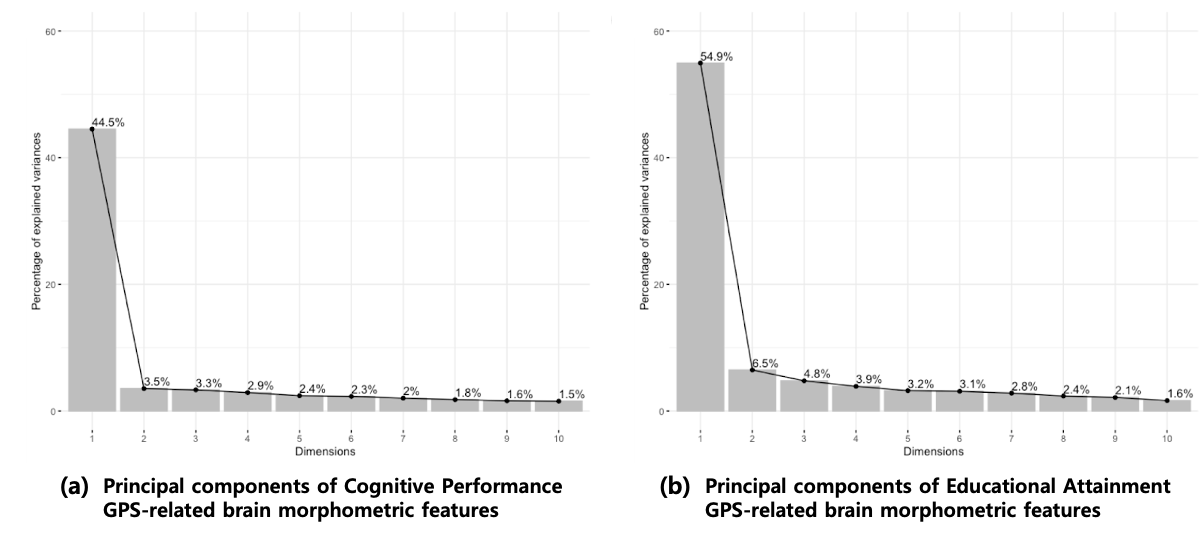

**Supplementary Figure 4. PCA biplot of cognitive GPSs-related brain morphometric features regarding covariates.**

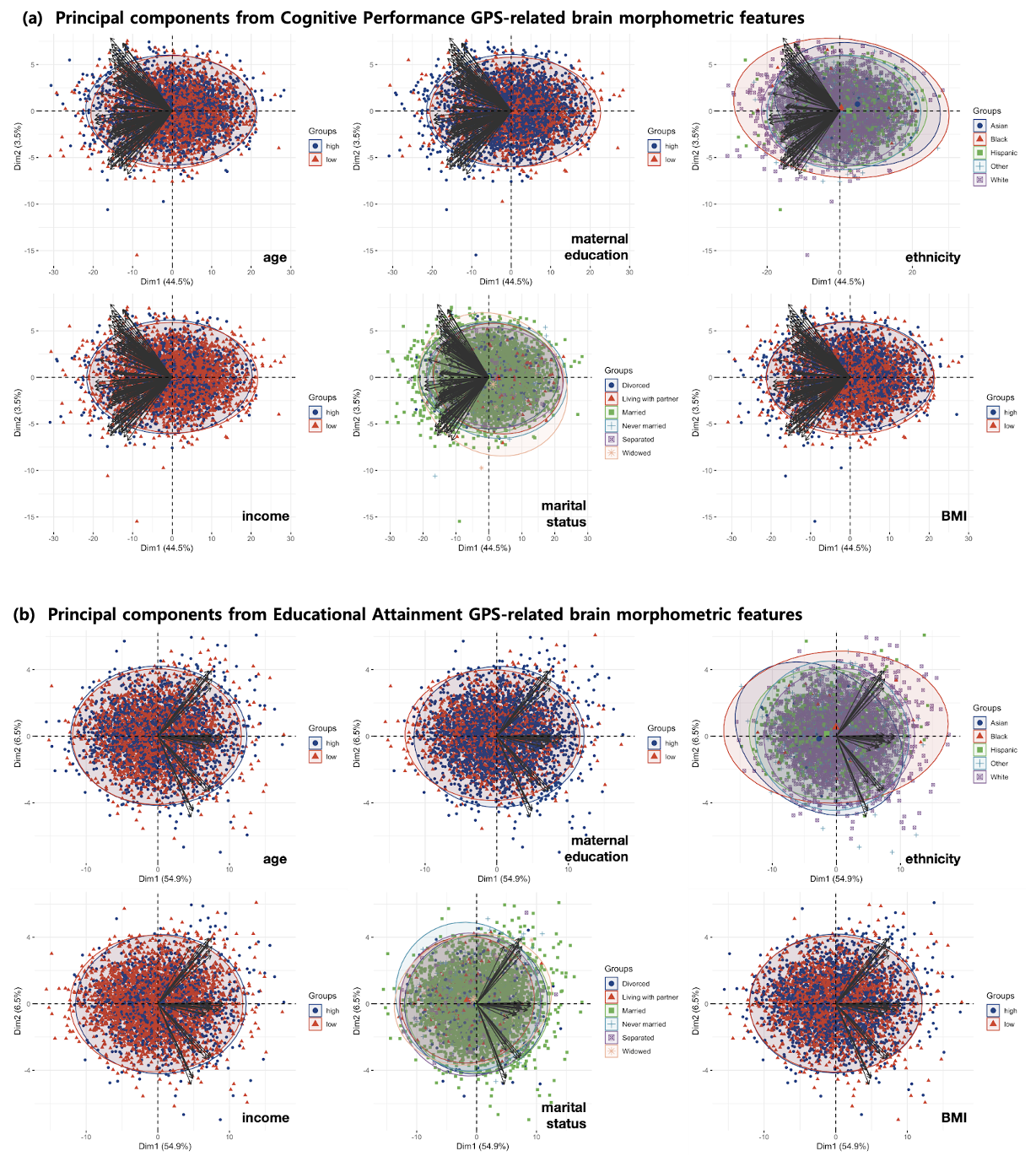

**Supplementary Figure 5.** **PCA biplot of cognitive GPSs-related brain morphometric features regarding sex.** The p-values from the Welch t-test were below 2.2e-16 both in Brain_CP GPS_ and Brain_EA GPS._

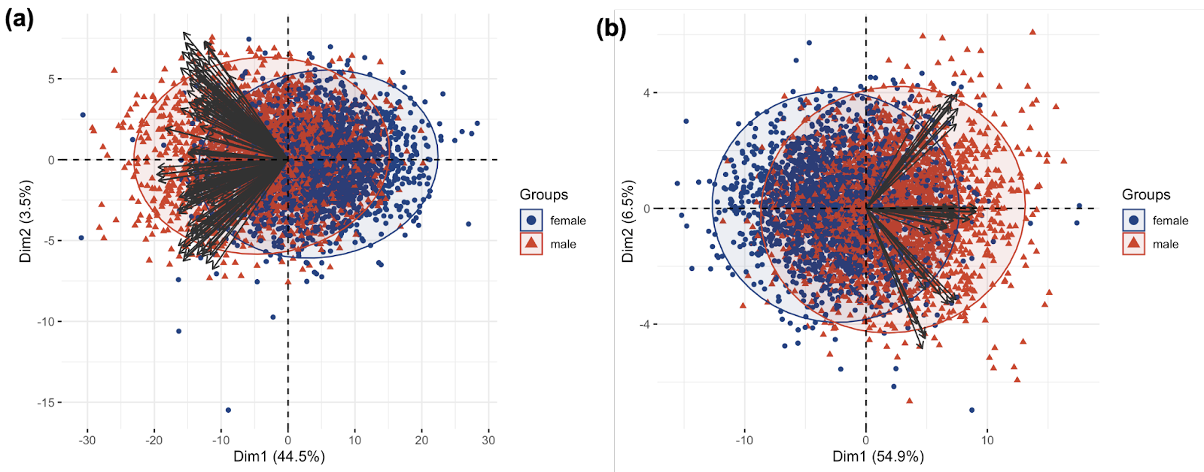

**Supplementary Figure 6. Effect of ELS on the relationships of CP GPS-brain-cognition.** The impact of *household challenges*, *neglect*, and *abuse* were not significantly moderate the effect of Cognitive Performance GPS on Brain_CP GPS_, consequently cognition.

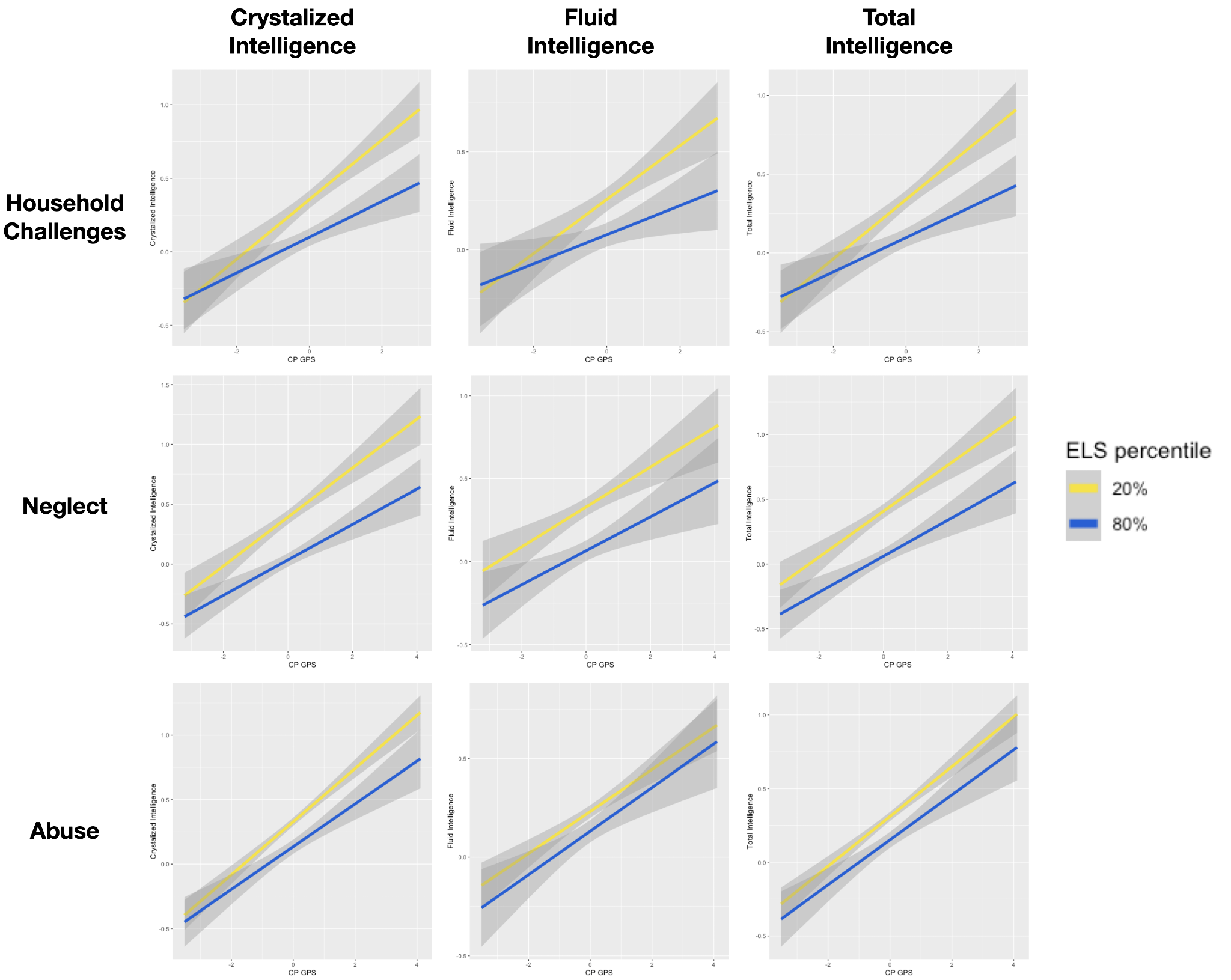

**Supplementary Figure 7. Effect of *household challenges* and *neglect* on the relationships of EA GPS-brain-cognition.** The impact of *household challenges* and *abuse* were not significantly moderate the effect of Educational Attainment GPS on Brain_EA GPS_, consequently cognition.

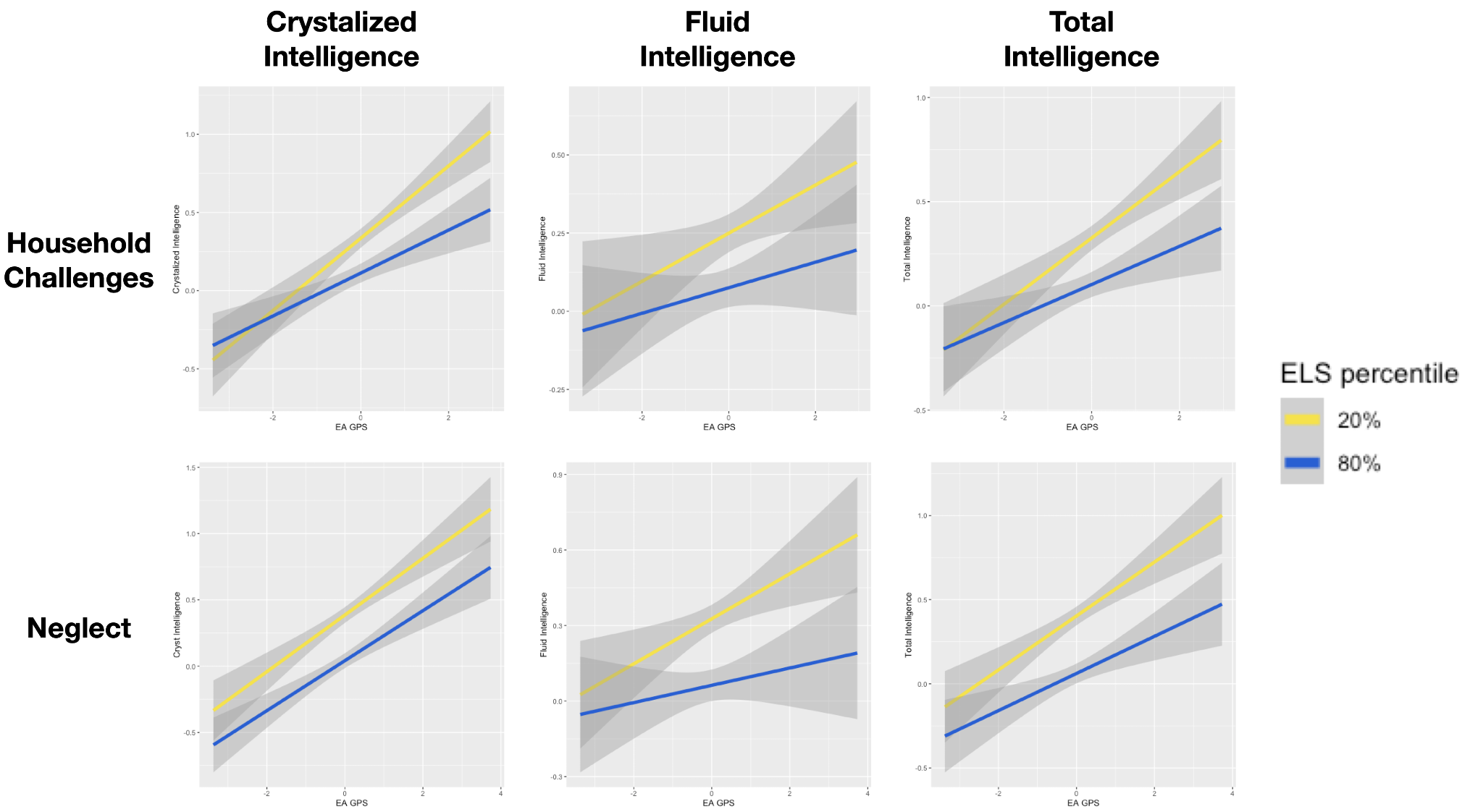
